# Effect of everyday discrimination on depression and suicidal ideation during the COVID-19 pandemic: a large-scale, repeated-measures study in the *All of Us* Research Program

**DOI:** 10.1101/2021.12.06.21266524

**Authors:** Younga H. Lee, Zhaowen Liu, Daniel Fatori, Joshua R. Bauermeister, Rebecca A. Luh, Cheryl R. Clark, Sarah Bauermeister, André R. Brunoni, Jordan W. Smoller

**Author notes:** **Correspondence:** Jordan W. Smoller, MD, ScD, Mailing Address: 185 Cambridge St, 6th Fl, Boston, MA 02114.

## Abstract

**Objective:** The COVID-19 pandemic has coincided with an increase in depressive symptoms as well as a growing awareness of health inequities and structural racism in the United States. Here, we examine the mental health impact of everyday discrimination during the pandemic in a large and diverse cohort of the *All of Us* Research Program.

**Methods:** Using repeated assessments of 62,651 participants in May to July of 2020, we fitted mixed-effects models to assess the effect of everyday discrimination on moderate to severe depression (Patient Health Questionnaire (PHQ)-9 ≥ 10) and suicidal ideation (PHQ-9 item 9 > 0), and applied inverse probability weights to account for non-random probabilities of completing the voluntary survey.

**Results:** Everyday discrimination was associated with increased odds of depression (adjusted odds ratio (aOR) [95% CI]: 1.21 [1.20 -1.22]) and suicidal ideation (1.17 [1.16-1.18]). For depression, the effects were larger in earlier phases of the pandemic (interaction p=8.2×10^−5^), which varied by main reason for discrimination and self-reported race and ethnicity. Among those who identified race or ancestry/national origin(s) as a primary reason for discrimination, Asian and Black or African American participants had 24% and 17% increase in the odds of depression in May of 2020 (1.24 [1.17-1.31] and 1.17 [1.12-1.22]), respectively, versus a 3% and 7% increase in July (1.03 [0.96-1.10] and 1.07 [1.02-1.12]).

**Conclusion:** In this large and diverse sample, increased levels of everyday discrimination were associated with higher odds of depression, particularly during the early phase of the pandemic among participants self-identifying as Asian or Black.

## Introduction

The COVID-19 pandemic continues to have far-reaching health and economic consequences for the American public, particularly for communities that face structural racism—such as Asian, Black, Hispanic or Latino, American Indian and Alaska Native, and Pacific Islander populations (1,2). During the pandemic, these communities have experienced higher rates of unemployment, food and housing insecurity than other groups (3,4). These challenges were compounded by the social, political, and economic trauma experienced at the height of the pandemic (5,6).

An increase in racially-motivated attacks targeting Asian and Pacific Islander individuals has been reported across the United States; this may be related to the ethnically-biased misrepresentations of the origins of COVID-19 across social media platforms (7). For example, studies have reported an increase in online hatred and racially-motivated posts towards Asian American communities after the former president of the U.S. referred to the coronavirus as the “Chinese virus” in March of 2020 (8–10). At the same time, there were more than 9,000 anti-Asian incidents reported in March to June of 2020 (11). In a 2020 survey by the Pew Research Center, Asian American respondents were more likely to report adverse experiences owing to their race or ethnicity since the coronavirus outbreak (6).

Prior to the pandemic, African American individuals were already experiencing limited health insurance coverage, reduced access to healthcare, and poor health outcomes (12,13). These long-standing health inequities have been further amplified over the course of the pandemic in association with higher rates of COVID-19 infection, hospitalization, and death (14,15), coupled with a sharp rise in unemployment and wage loss due to the pandemic (16). Furthermore, the killing of Black men and women including George Floyd and Breonna Taylor spotlighted structural racism, systemic inequities, and experiences of discrimination due to race and ethnicity (17).

An individual’s experience of discrimination has been reported to have an adverse impact on both physical and mental health among Asian, Black, and Hispanic or Latino adults (18–21). When comparing across nine ethnic subgroups in the United States, Afro-Caribbean American participants (including Haitian, Jamaican, and Trinidadian/Tobagonian) reported highest rates of discrimination (22). However, Asian (including Vietnamese, Filipino, and Chinese) and Latino American (including Cuban, Portuguese, and Mexican) participants were most likely to experience harmful psychological consequences in response to discrimination.

In the present study, we hypothesized that the likelihood of having moderate to severe depression and/or suicidal ideation would be associated with increasing levels of everyday discrimination during the COVID-19 pandemic. We additionally examined factors that may modify this relationship—such as the main reason for discrimination, survey timing, self-reported race and ethnicity, and pre-pandemic mood disorder diagnosis. Lastly, we investigated the effects of discrimination specifically due to race or ancestry/national origin(s) on depression and potential variations by survey timing within and across four subgroups defined by self-reported race and ethnicity.

## Methods

### Cohort description

To date, the *All of Us* Research Program (hereafter, AoU) has enrolled more than 400,000 participants, of which more than 80% are individuals from communities that have been underrepresented in biomedical research, such as people identifying as Asian, Black, Hispanic or Latino race and ethnicity, LGBTQ+ sexual orientation and gender identities, and those with less than high school education or household incomes at the federal poverty level (23). The diversity of the AoU cohort provides a unique opportunity to examine the impact of discrimination on mental health outcomes that varies across sociodemographic and environmental contexts (24). The Institutional Review Boards of the *All of Us* Research Program approved all study procedures, and all participants provided informed consent to share electronic health records (EHRs), surveys, and other study data with qualified investigators for broad-based research.

### Study sample

The <u>CO</u>VID-19 <u>P</u>articipant <u>E</u>xperience (COPE) survey is a brief online survey administered to the AoU participants to seek new insights into whether and how the pandemic affects people differently over time, starting in May of 2020 (25). The survey includes questions on COVID-19 symptoms, physical and mental health, social distancing, economic impacts, and coping strategies. At the time of analysis, there were 62,651 participants who had completed at least one of the three COPE surveys administered in May, June, or July of 2020, resulting in an overall participation rate of 19.9% (May: 13.6%, June: 10.4%, July: 9.2%). After excluding 3,789 participants with missing information on self-reported race and ethnicity, and 1,761 participants with missing information on the primary exposure, we then fit mixed-effects logistic regression models using 97,013 COPE surveys completed by 57,101 participants remaining in the study sample.

### Exposure

The primary exposure was the level of everyday discrimination measured using a revised version of the Everyday Discrimination Scale (18), a nine-item checklist that measures experiences of discrimination occurring in the past month, using a four-point Likert scale response format (ranging from *1 = Never* to *4 = Almost every day*). Discrimination was measured using the self-reported frequency of being treated with less courtesy, less respect, poor service, and being considered as dishonest, threatening, and not smart. The scale is scored by summing the responses across all items, thus resulting in total scores ranging from 9 to 36.

At the end of the Everyday Discrimination Scale, there is a follow-up question asking: *“What do you think is the main reason for these experiences? Select all that apply*.*”* We conducted a secondary analysis to examine the unique impact of discrimination specifically due to race or ancestry/national origin(s) on depression and suicidal ideation during the COVID-19 pandemic. In this analysis, we excluded 25,303 participants who reported discrimination solely due to other reasons than race or ancestry/national origin(s). We then fit mixed-effects logistic regression models using 53,619 COPE surveys completed by 35,021 participants remaining in the study sample.

### Outcome

Depression symptoms were assessed with the Patient Health Questionnaire (PHQ)-9 (26), a nine-item checklist that scores each of the nine DSM-IV criteria for depression in the past two weeks, using a four-point Likert scale response format (ranging from *0 = Not at all* to *3 = Nearly every day*). The scale is scored by summing their responses to all nine items, with total scores ranging from 0 to 27; total scores of 5, 10, 15, and 20 represent cut-points for mild, moderate, moderately severe, and severe depression (26,27). Here, we refer to scores of 10 or above as “moderate to severe depression.”

In addition to the total score, we separately examined the ninth item of PHQ-9, which evaluates passive thoughts of death or self-injury within the last two weeks, as a secondary outcome. This item is often used to screen depressed patients for suicide risk using the following question: *“In the last two weeks, how often have you thought that you would be better off dead or of hurting yourself in some way?”* As with the other questions in the PHQ-9, responses are based on the same four-point Likert scale for frequency, and any non-zero response was considered as positive for suicidal ideation.

### Inverse probability weighting to adjust for potential selection bias

It has been previously reported that research volunteers tend to be healthier and have higher socioeconomic status compared to the underlying general population (28). To address this possible “healthy volunteer bias” among the COPE respondents, we first compared the distribution of sociodemographic covariates between our study sample against the overall AoU cohort. We calculated predicted probabilities of survey completion for each month by fitting three separate logistic regression models with sex assigned at birth, self-reported race and ethnicity, birthplace, college education, marital/partnership status, health insurance status, employment status, homeownership, and current age as predictors. We then took the inverse of the product of predicted probabilities to derive weights for mixed-effects models.

### Extraction of pre-pandemic mood disorder diagnosis

We used linked EHRs to establish whether respondents had a pre-pandemic history of mood disorder. Participants were considered as cases of pre-pandemic mood disorder if they had two or more qualifying diagnostic codes (identified based on Systematized Nomenclature of Medicine (SNOMED) code 46206005) within one year prior to January 21^st^, 2020—the date of the first reported COVID-19 case in the United States (29). Of 62,551 participants in the study sample,11,075 (17.7%) had linked EHRs, and those who did not meet the criteria or, to be conservative, did not have linked EHRs were considered as non-cases.

### Analysis of repeated-measures data

As noted, the survey was administered to participants three times during the early phase of the pandemic, though not all respondents completed each monthly survey. To accommodate both missing responses and the correlated nature of repeated measurements (30), we fitted mixed-effects logistic regression models using the *lme4* R package to determine the relationships between the repeated survey measures with subject-specific random intercepts and fixed effects for the timing of survey administration (e.g., May, June, and July of 2020). In addition to estimating odds ratios from these models, we used the *ggpredict* function from the *ggeffects* R package to obtain and visualize predicted probabilities of having moderate to severe depression estimated at different levels of discrimination.

We adjusted for potential confounding factors including sex assigned at birth, current age, homeownership, employment status, educational attainment, health insurance status, and self-reported race and ethnicity (except in analyses stratified by self-reported race and ethnicity). We further adjusted the mixed-effects models for the experience of COVID-19 symptoms for each survey timing (measured as part of the COPE survey), given the possibility that COVID-19 infection could be a confounding factor in the relationship between discrimination (31) and depression (32). Lastly, we tested for variations by survey timing, pre-pandemic mood disorder diagnosis, and self-reported race to explore factors that potentially modify the effects of discrimination on depression or suicidal ideation.

All analyses were performed using data from the *All of Us Registered Tier Version R2020Q4R2* on the AoU Researcher Workbench, a cloud-based platform where approved researchers can access and analyze data (23). We used R version 4.1.0 in a Jupyter Notebook contained in the AoU Workbench to query data, perform statistical analyses, and generate tables and figures. In all analyses, a two-tailed value of *p<*0.05 was considered statistically significant.

## Results

### Descriptive analysis

As shown in the first two columns of Table 1, participants in the study sample (N = 62,651) were more likely to be female (65.6 vs. 60.7%), White (82.3 vs. 53.0%), and non-Hispanic (93.7 vs. 81.9%) compared to the underlying AoU population (N = 314,994). They were also more likely to have received a college education or beyond, own a home, have a partner, be older, and have higher annual household income. In addition, the prevalence of pre-pandemic mood disorder was lower in the study sample compared to the underlying AoU cohort (5.6 vs. 6.5%). Overall, then, participants selected into our study sample were less likely to experience socioeconomic deprivation and more likely to be healthy relative to the underlying AoU cohort—a potential indication of healthy volunteer bias commonly observed in volunteer-based cohorts (33). We addressed this issue statistically by applying inverse probability weights to the mixed-effects models.

**Table 1.** Sociodemographic characteristics of the study sample and a comparison against the underlying *All of Us* Research Program cohort (hereafter, AoU cohort).

|  | Overall<br>AoU cohort |  | Reported<br>discrimination due<br>to any reason(s) |  | Reported<br>discrimination<br>due to race or<br>ancestry/national<br>origin(s) |  |
| --- | --- | --- | --- | --- | --- | --- |
|  | (N=314,994) |  | (N=62,651) |  | (N=37,348) |  |
| *Categorical variables* | N | % | N | % | N | % |
| Sex assigned at birth |  |  |  |  |  |  |
| Male | 119,750 | 38.0 | 21,158 | 33.8 | 13,471 | 36.1 |
| Female | 191,114 | 60.7 | 41,084 | 65.6 | 23,641 | 63.3 |
| Not male, not female/Prefer not to answer,<br>or skipped/No matching concept | 4,130 | 1.3 | 409 | 0.7 | 236 | 0.6 |
| Self-reported race and ethnicity |  |  |  |  |  |  |
| Asian | 10,562 | 3.4 | 1,813 | 2.9 | 1,243 | 3.3 |
| Another single population/None of these<sup>†</sup> | 5,635 | 1.8 | 783 | 1.2 | 447 | 1.2 |
| Black or African American | 68,112 | 21.6 | 3,570 | 5.7 | 2,176 | 5.8 |
| More than one population<sup>†</sup> | 5,619 | 1.8 | 1,104 | 1.8 | 632 | 1.7 |
| White | 166,917 | 53.0 | 51,592 | 82.3 | 30,523 | 81.7 |
| Missing | 58,149 | 18.5 | 3,789 | 6.0 | 2,327 | 6.2 |
| Hispanic | 57,005 | 18.1 | 3,958 | 6.3 | 2,398 | 6.4 |
| Born in the United States | 262,791 | 83.4 | 56,645 | 90.4 | 33,331 | 89.2 |
| College educated or beyond | 211,572 | 67.2 | 57,476 | 91.7 | 34,602 | 92.6 |
| Employed | 149,504 | 47.5 | 34,195 | 54.6 | 18,900 | 50.6 |
| Insured | 283,089 | 89.9 | 61,039 | 97.4 | 36,536 | 97.8 |
| Homeowner | 136,530 | 43.3 | 43,583 | 69.6 | 27,553 | 73.8 |
| Married/partnered | 150,766 | 47.9 | 39,723 | 63.4 | 24,648 | 66.0 |
| Had pre-pandemic mood disorder dx | 20,560 | 6.5 | 3,525 | 5.6 | 1,712 | 4.6 |
| *Continuous variables* | Mean | SD | Mean | SD | Mean | SD |
| Current age (in years) | 53.9 | 16.7 | 58.9 | 15.9 | 61.4 | 15.4 |
| Annual household income (in dollars) | 61,410.7 | 54,136.7 | 91,549.2 | 59,339.5 | 95,594.6 | 59,297.3 |
| Number of co-resident(s) | 1.7 | 1.6 | 1.4 | 1.3 | 1.4 | 1.2 |

**Abbreviations:** SD, standard deviation; dx, diagnosis.
<sup>†</sup> ‘More than one population’ and ‘Another single population/None of these’ were collapsed in the analysis due to limited sample size. The missing category included participants whose responses were either “I prefer not to answer”, “None indicated”, or “Skipped.”

In addition, the distribution of total score and the main reason for discrimination varied substantially by survey timing and self-reported race and ethnicity. Black or African American participants reported the highest levels of discrimination, especially in June of 2020 (mean [SD]: 5.96 [4.78]), followed by participants self-identifying as Asian and those self-identifying as either “More than one population,” “Another single population,” or “None of these” (see Supplemental Figures 1a-b). In addition, there were more substantial variations with the main reason for discrimination. While age and gender were most frequently reported as the main reason for discrimination by White participants, ancestry or national origin and race were most frequently reported by Asian and Black or African American participants, respectively.

Lastly, the overall prevalence of moderate to severe depression was highest in May of 2020 for a prevalence of 16.3%, which decreased up to 1.4% in the following months (see Supplemental Table 1). Of 7,142 cases of moderate to severe depression identified in May of 2020, 59.8% of them had moderate depression, 26.6% had moderately severe depression, and 13.6% had severe depression. The overall prevalence of suicidal ideation was 5.9% in May of 2020, and remained relatively stable in the subsequent months.

### Mixed-effects logistic regression analysis

We observed significantly increased odds of moderate to severe depression and suicidal ideation as the levels of everyday discrimination increased. On average, participants experienced a 21% (adjusted odds ratio (aOR) [95% CI]: 1.21 [1.20-1.22]) increase in the odds of having moderate to severe depression and a 17% (1.17 [1.16-1.18]) increase in odds of suicidal ideation for each one-point increase in everyday discrimination (see Table 2 for odds ratios and Supplemental Figure 2a-d for predicted probabilities). After the addition of a discrimination-by-survey timing interaction term, we saw an attenuation of the discrimination-depression association over the course of the survey period (interaction *p*=2.0×10^−4^) (see Supplemental Figure 3a-b). Strikingly, the time-varying nature of this association was more evident if the main reason was due to race or ancestry/national origin(s) (interaction *p*=4.5×10^−5^).

**Table 2.** Mixed-effects logistic regression analysis examining the effects of everyday discrimination on moderate to severe depression and suicidality with variations by survey timing, self-reported race and ethnicity, and pre-pandemic mood disorder diagnosis.

| Model configuration | Main reason for discrimination | Stratified results | Clinically elevated depression (PHQ-9 total score $\geq 10$) | | | Suicidality (PHQ-9 item 9 > 0) | | |
| --- | --- | --- | --- | --- | --- | --- | --- | --- |
|  |  |  | aOR | 95% CI | p-value | aOR | 95% CI | p-value |
| Base model | Any | Overall | 1.21 | (1.20 - 1.22) | <2.0E-16 | 1.17 | (1.16 - 1.18) | <2.0E-16 |
|  | Race or ancestry/national origin(s) | Overall | 1.17 | (1.15 - 1.19) | <2.0E-16 | 1.15 | (1.13 - 1.18) | <2.0E-16 |
| Base model + an interaction with survey timing | Any | Stratified - May 2020 | 1.17 | (1.16 - 1.19) | 8.2E-05<sup>†</sup> | 1.18 | (1.18 - 1.19) | 4.9E-01<sup>†</sup> |
|  |  | Stratified - June 2020 | 1.17 | (1.15 - 1.19) |  | 1.17 | (1.16 - 1.18) |  |
|  |  | Stratified - July 2020 | 1.16 | (1.15 - 1.18) |  | 1.17 | (1.16 - 1.18) |  |
|  | Race or ancestry/national origin(s) | Stratified - May 2020 | 1.16 | (1.15 - 1.18) | 4.5E-05<sup>†</sup> | 1.13 | (1.11 - 1.15) | 8.2E-01<sup>†</sup> |
|  |  | Stratified - June 2020 | 1.13 | (1.11 - 1.14) |  | 1.10 | (1.08 - 1.12) |  |
|  |  | Stratified - July 2020 | 1.13 | (1.11 - 1.14) |  | 1.10 | (1.08 - 1.12) |  |
| Base model + an interaction with pre-pandemic mood disorder dx | Any | Stratified - Without dx | 1.21 | (1.20 - 1.22) | 2.0E-04<sup>†</sup> | 1.17 | (1.16 - 1.18) | 1.3E-01<sup>†</sup> |
|  |  | Stratified - With dx | 1.16 | (1.13 - 1.19) |  | 1.16 | (1.13 - 1.19) |  |
|  | Race or ancestry/national origin(s) | Stratified - Without dx | 1.18 | (1.16 - 1.20) | 1.9E-01<sup>†</sup> | 1.16 | (1.13 - 1.19) | 5.4E-02<sup>†</sup> |
|  |  | Stratified - With dx | 1.11 | (1.05 - 1.17) |  | 1.10 | (1.03 - 1.16) |  |

**Abbreviations:** aOR, adjusted odds ratio; CI, confidence interval; dx, diagnosis.
**Note:** All models were adjusted for sex assigned at birth, self-reported race and ethnicity, current age, homeownership, employment status, educational attainment, health insurance status, experience of COVID symptom(s), and diagnosis of mood disorder(s) within one year prior to the COVID-19 pandemic.
<sup>†</sup> Indicates p-values for the interaction term being tested in a given model.

Interestingly, the sign of a discrimination-by-pre-pandemic mood disorder diagnosis interaction was also negative (interaction *p*=2.0×10^−4^). While the presence of a pre-pandemic mood disorder was associated with higher predicted probabilities of moderate to severe depression at lower levels of discrimination, participants without a pre-existing mood disorder diagnosis had similarly elevated probability of depression upon exposure to higher levels of discrimination (see Figure 2). Subsequently, we conducted stratified analyses to estimate the main effect of discrimination and interaction with survey timing within and across subgroups defined based on self-reported race and ethnicity. We observed that the odds of depression were highest among Asian and Black or African American participants early in the pandemic (May and June), particularly when the main reason for discrimination was due to race or ancestry/national origin(s) (interaction p=6.4×10^−12^ and 7.9×10^−3^, respectively; see Table 3 and Figure 1b).

**Table 3.**
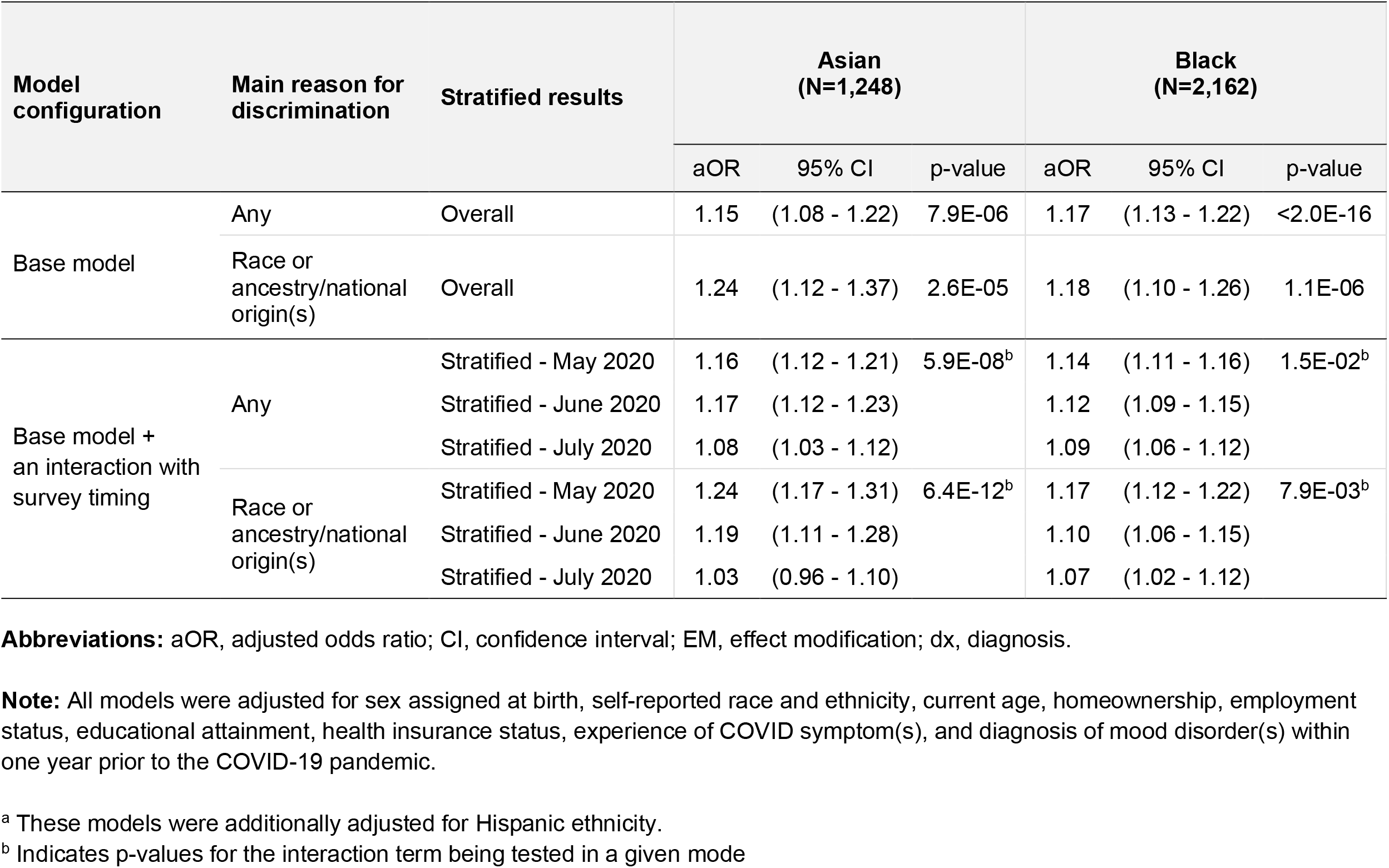
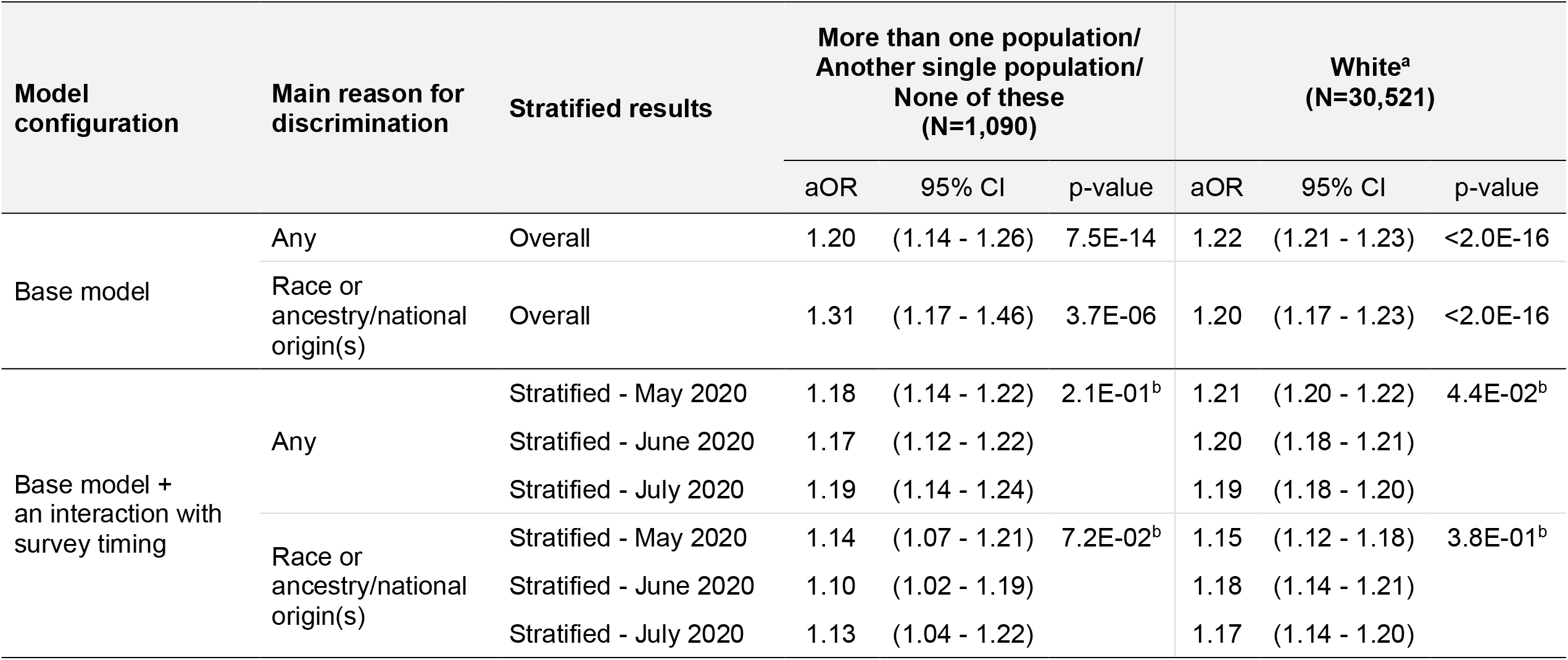
Stratified mixed-effects logistic regression analysis examining the effects of everyday discrimination on moderate to severe depression (PHQ-9 total score ≥ 10) by self-reported race and ethnicity.

**Figure 1.**
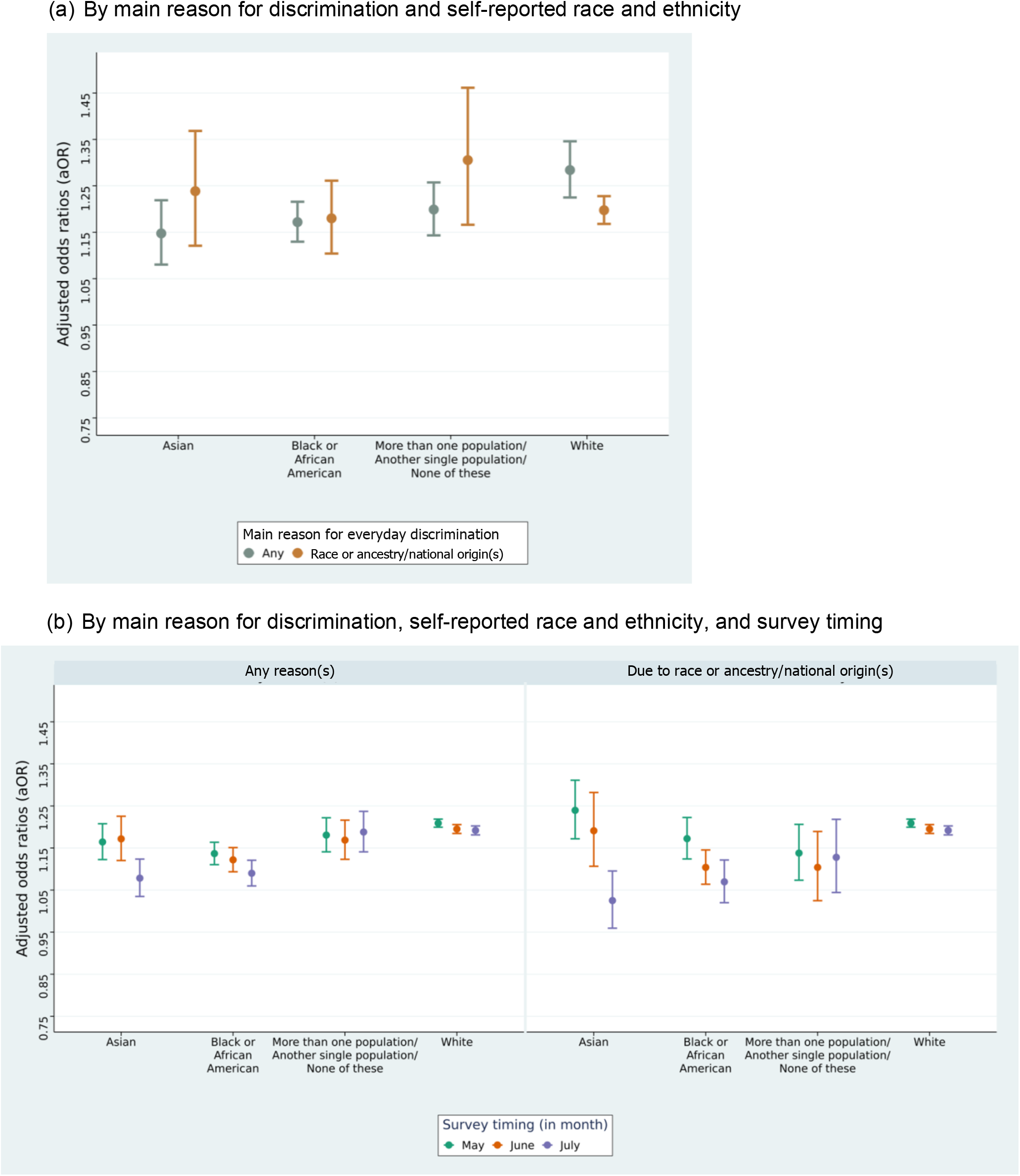
Variations in the effects of everyday discrimination on moderate to severe depression by main reason for everyday discrimination, self-reported race and ethnicity, and survey timing.

**Figure 2.**
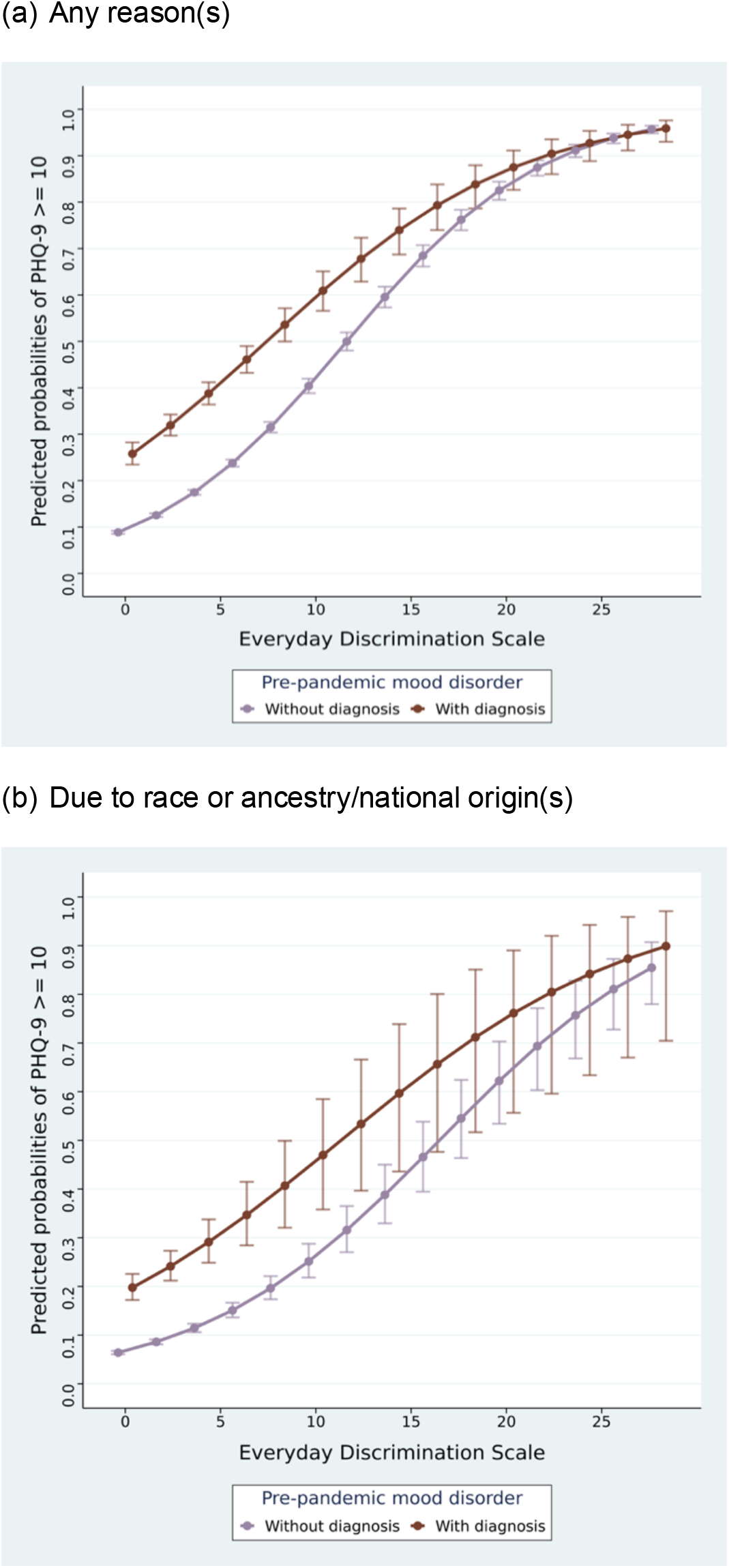
Variations in the effects of everyday discrimination on moderate to severe depression (PHQ-9 total score ≥ 10) by pre-pandemic mood disorder diagnosis.

## Discussion

Everyday discrimination has been implicated as a strong risk factor for adverse mental health outcomes. We were able to leverage a large and diverse cohort, derived from the *All of Us* Research Program, to examine the longitudinal effect of discrimination on depressive symptoms and suicidal ideation and multiple factors potentially modifying this relationship. First, we observed significant effects of discrimination on moderate to severe depression and suicidal ideation (measured using the PHQ-9) during the COVID-19 pandemic. Next, stratified analyses in subgroups defined by self-reported race and ethnicity shed light on variations by the main reason for discrimination and survey timing. Notably, the odds of depression were highest among Asian and Black or African American participants early in the pandemic (May and June of 2020), particularly when the main reason for discrimination was due to race or ancestry/national origin(s).

Our study adds to prior work implicating discrimination as a risk factor for adverse mental health outcomes. Lee and Waters (34) examined the effects of significant racial discrimination on mental, physical, and sleep health problems during first months of the pandemic in a sample of 410 adults self-identifying as Asian and currently residing in the United States. The participants experienced a nearly 30% increase in racial discrimination, which was experienced in various forms (e.g., hate crimes, microaggressions, and vicarious discrimination) and had particularly strong associations with anxiety and depressive symptoms. In a larger cross-sectional study that included both Asian and Black American adults, Chae and colleagues (35) focused specifically on experiences of vicarious racism—defined as hearing about or seeing racists acts directed against members of one’s racial group—and vigilance about racial discrimination. In that study, increasing exposure to either vicarious racism and racial discrimination vigilance was associated with more symptoms of depression and anxiety, although the study did not assess clinically significant depression.

Wu and colleagues (36) examined COVID-19-related discrimination using repeated measurements from Asian and White individuals (N = 7,778) who participated in over 13 waves of an internet panel tracking survey between March 10^th^, 2020 and September 30^th^, 2020. They further distinguished Asian participants who were born in the United States from those born outside the United States. Relative to White participants, both Asian American and Asian immigrant participants were more likely to report depressive symptoms (measured using the PHQ-4 scale). Within the Asian sample, Asian immigrant participants reported higher levels of acute discrimination in March of 2020, whereas Asian American participants reported significantly higher levels of discrimination in April and early May of 2020.

Our study has several strengths relative to prior reports. To our knowledge, this is the largest and most diverse study conducted in the United States examining the mental health impact of everyday discrimination during the COVID-19 pandemic. We leveraged repeated measurements of discrimination and depression and examined temporal fluctuations within and across four subgroups defined based on self-reported race and ethnicity. We further included COVID-19 symptoms as a covariate in our adjusted models to remove potential confounding effects of COVID-19 infection on mental health outcomes. In addition, because the COPE respondent sample may not have been fully representative of the larger *All of Us* cohort, we applied inverse probability weighting to minimize selection bias. Lastly, we leveraged linked EHRs to examine potential effect modification of the impact of discrimination by pre-existing mood disorder diagnosis.

Our study should also be evaluated in light of several limitations. First, the first COPE survey was administered in May of 2020, after the start of the pandemic. Since we did not have pre-pandemic measurements of discrimination or PHQ-9 scores, we could not capture the evolution of their relationship at the beginning of the pandemic. Second, we also could not establish a causal relationship between discrimination and depression because these were measured concurrently during waves of the survey. Third, while our use of inverse probability weighting should address discrepancies between COPE respondents and the overall AoU cohort, the latter is not a fully representative sample of the U.S. population; thus, the generalizability of these findings warrants caution.

## Conclusion

In a large-scale, longitudinal investigation of the impact of everyday discrimination on mental health outcomes during the COVID-19 pandemic, we find that discrimination was associated with both depressive symptoms and suicidal ideation. For those who reported discrimination primarily due to race, ancestry, or national origin, the effect was greater in May and June of 2020, especially among Asian and Black or African American respondents. In addition, at high levels of discrimination, the likelihood of depression was similar among those with or without a pre-pandemic mood disorder diagnosis. These results illustrate the complex and dynamic relationship between discrimination and adverse mental health outcomes.

## Data Availability

De-identified data are available on the Researcher Workbench of the *All of Us* Research Program located at https://workbench.researchallofus.org.

## Acknowledgments

The *All of Us* Research Program is supported by grants through the National Institutes of Health, Office of the Director: Regional Medical Centers: 1 OT2 OD026549; 1 OT2 OD026554; 1 OT2 OD026557; 1 OT2 OD026556; 1 OT2 OD026550; 1 OT2 OD 026552; 1 OT2 OD026553; 1 OT2 OD026548; 1 OT2 OD026551; 1 OT2 OD026555; IAA#: AOD 16037; Federally Qualified Health Centers: HHSN 263201600085U; Data and Research Center: 5 U2C OD023196; Biobank: 1 U24 OD023121; The Participant Center: U24 OD023176; Participant Technology Systems Center: 1 U24 OD023163; Communications and Engagement: 3 OT2 OD023205; 3 OT2 OD023206; and Community Partners: 1 OT2 OD025277; 3 OT2 OD025315; 1 OT2 OD025337; 1 OT2 OD025276 In addition to the funded partners, the *All of Us Research Program* would not be possible without the contributions made by its participants.

All authors are supported by the International HundredK+ Cohorts Consortium (IHCC), which has been created in collaboration with the Global Alliance for Genomics and Health (GA4GH) and the Global Genomics Medicine Collaborative (G2MC) with support from the National Institute of Health and the Wellcome Trust. In addition, J.R.B. and S.B. are supported by: Dementias Platform UK (DPUK) funded by the Medical Research Council (MRC) MR/T0333771. J.W.S. is supported in part by a gift from the Demarest Lloyd, Jr. Foundation. Lastly, we thank Dr. Nhi-Ha Trinh at Massachusetts General Hospital for her valuable input to this manuscript.

