## Supplemental Table 1 for "Effect of everyday discrimination on depression and suicidal ideation during the COVID-19 pandemic: a large-scale, repeated-measures study in the *All of Us* Research Program"

**Supplemental Materials**

**Supplemental Table 1.** Distribution of depression severity by survey timing.

| Survey timing | Level of depression severity, PHQ-9 score | N | Prevalence (%) |
|---|---|---|---|
| May 2020 (N=43,903) | Minimal, 0–4 | 26,155 | 59.6 |
| | Mild, 5–9 | 10,606 | 24.2 |
| | Moderate, 10–14 | 4,272 | 9.7 |
| | Moderately severe, 15–19 | 1,898 | 4.3 |
| | Severe, 20–27 | 972 | 2.2 |
| | Moderate to severe, 10-27 | 7,142 | 16.3 |
| | Suicidal ideation (PHQ-9 item 9 > 0) | 2,603 | 5.9 |
| June 2020 (N=33,546) | Minimal, 0–4 | 20,817 | 62.1 |
| | Mild, 5–9 | 7,728 | 23.0 |
| | Moderate, 10–14 | 2,944 | 8.8 |
| | Moderately severe, 15–19 | 1,419 | 4.2 |
| | Severe, 20–27 | 638 | 1.9 |
| | Moderate to severe, 10-27 | 5,001 | 14.9 |
| | Suicidal ideation (PHQ-9 item 9 > 0) | 1,950 | 5.8 |
| July 2020 (N=29,794) | Minimal, 0–4 | 18,471 | 62.0 |
| | Mild, 5–9 | 6,794 | 22.8 |
| | Moderate, 10–14 | 2,681 | 9.0 |
| | Moderately severe, 15–19 | 1,242 | 4.2 |
| | Severe, 20–27 | 606 | 2.0 |
| | Moderate to severe, 10-27 | 4,529 | 15.2 |
| | Suicidal ideation (PHQ-9 item 9 > 0) | 1,717 | 5.7 |
