## Supplemental Figure 1 for "Effect of everyday discrimination on depression and suicidal ideation during the COVID-19 pandemic: a large-scale, repeated-measures study in the *All of Us* Research Program"

**Supplemental Figure 1.** Distribution of total scores on the Everyday Discrimination Scale among participants who reported any everyday discrimination.

(a) Total score on the Everyday Discrimination Scale

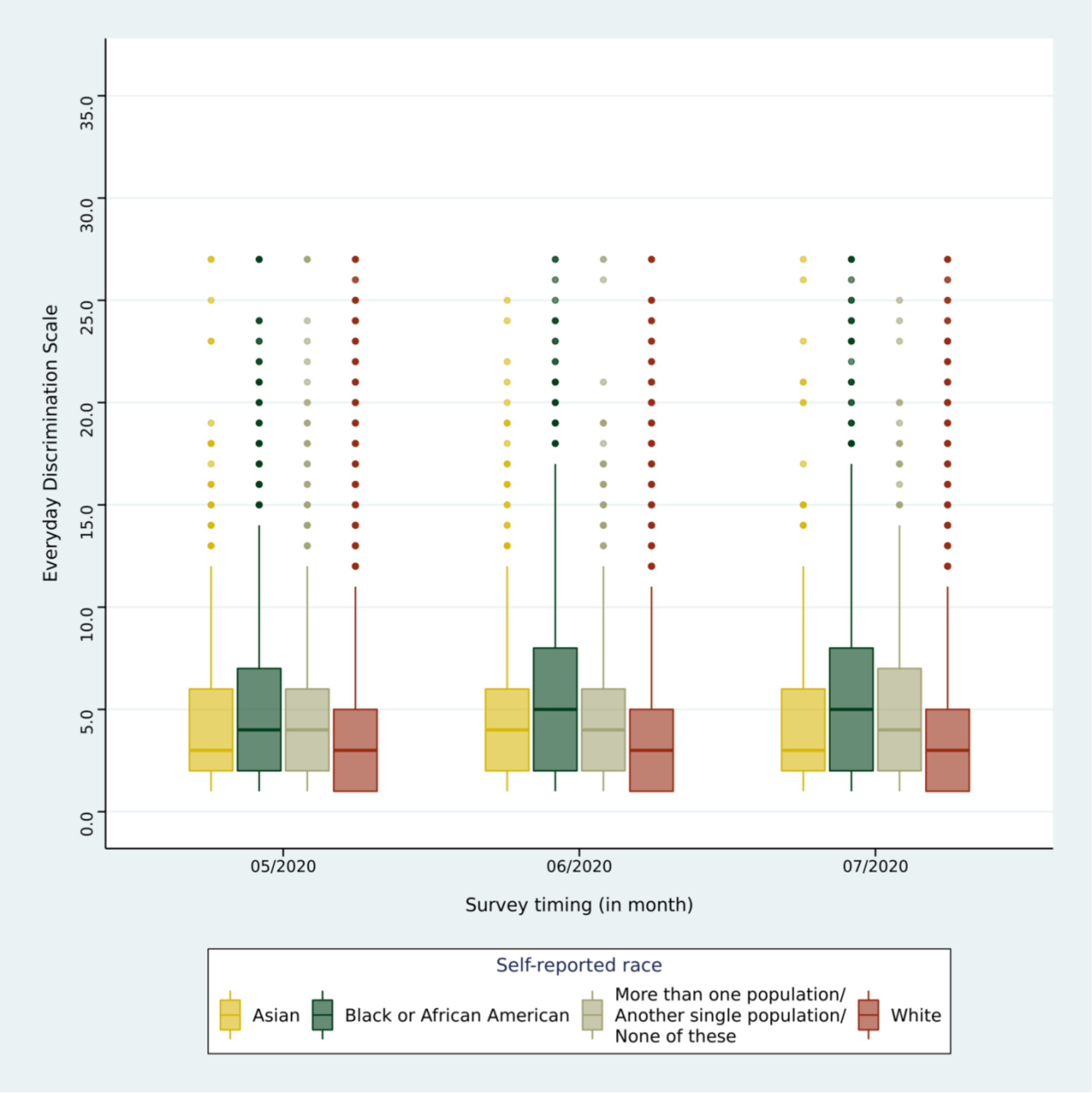

(b) Main reason(s) for everyday discrimination by survey timing and self-reported race and ethnicity.

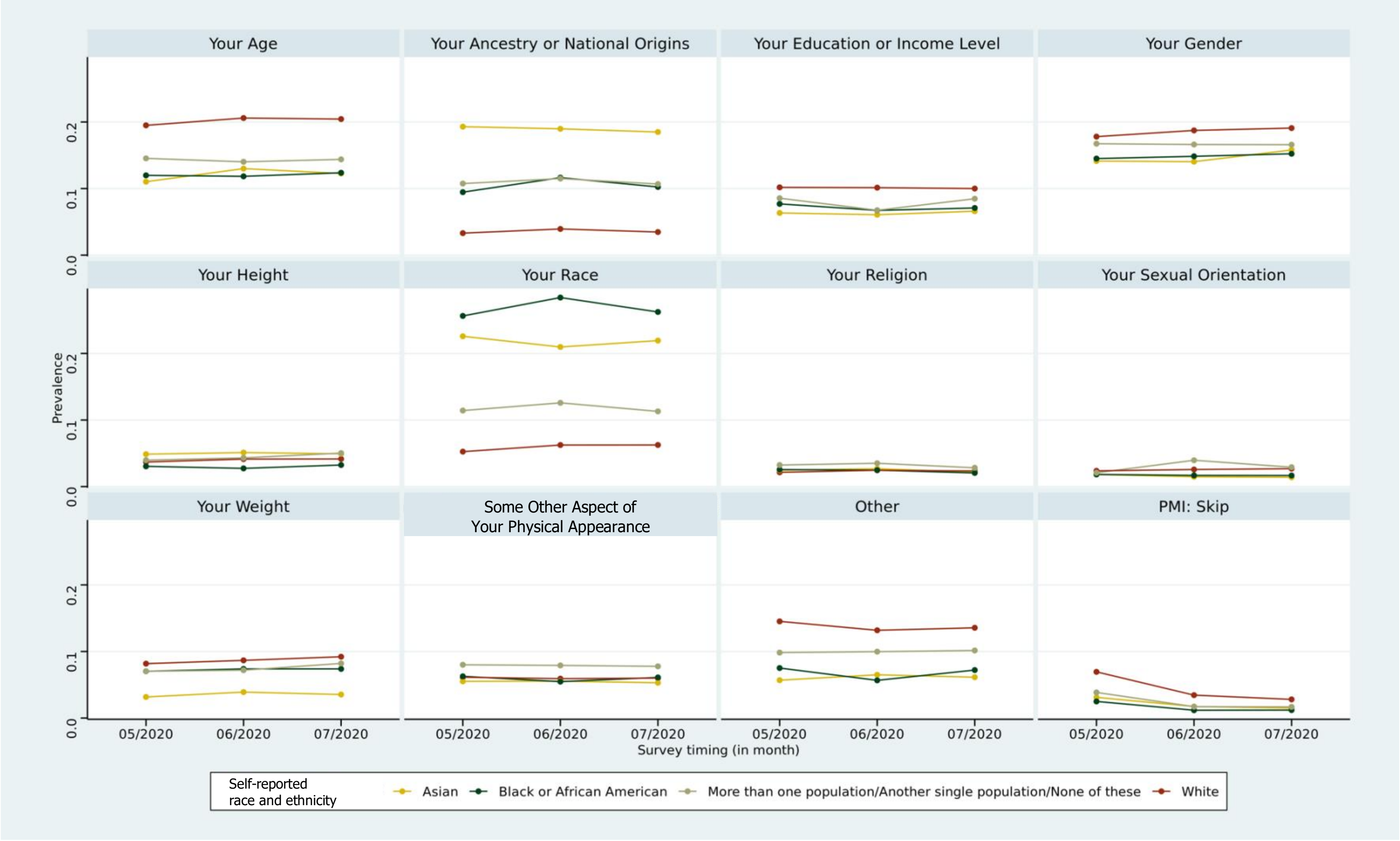
