## Supplemental Figure 2 for "Effect of everyday discrimination on depression and suicidal ideation during the COVID-19 pandemic: a large-scale, repeated-measures study in the *All of Us* Research Program"

**Supplemental Figure 2.** Overall effects of everyday discrimination on depressive symptoms and suicidal ideation.

<u>Outcome: Clinically elevated depression (PHQ-9 total score ≥ 10)</u>

(a) Any reason(s)

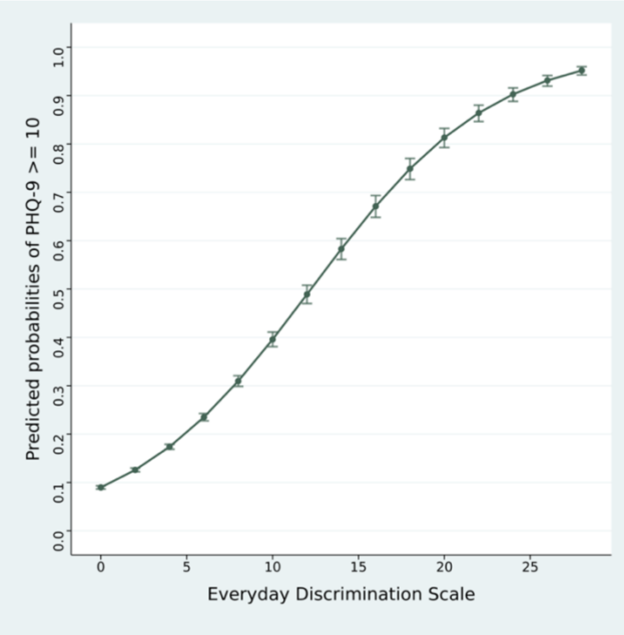


(b) Due to race or ancestry/national origin(s)

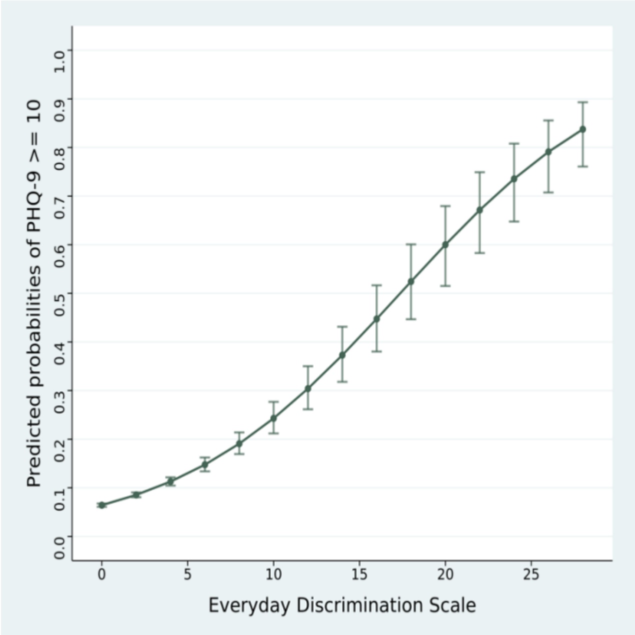


<u>Outcome: Suicidal ideation (PHQ-9 item 9 > 0)</u>

(c) Any reason(s)

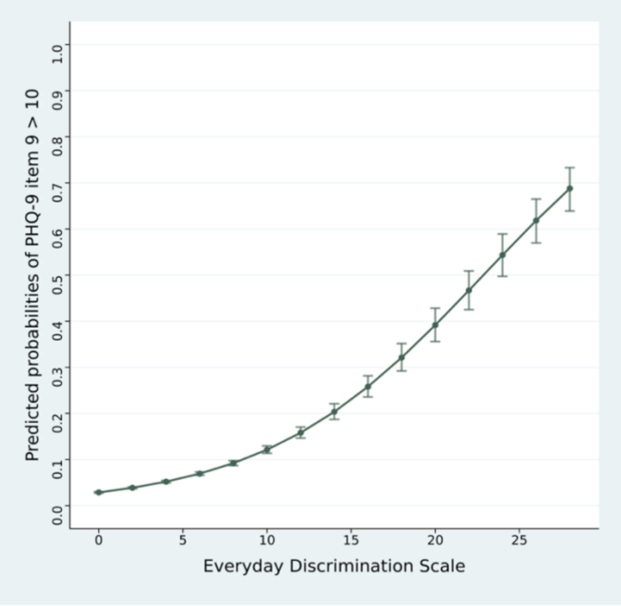


(d) Due to race or ancestry/national origin(s)

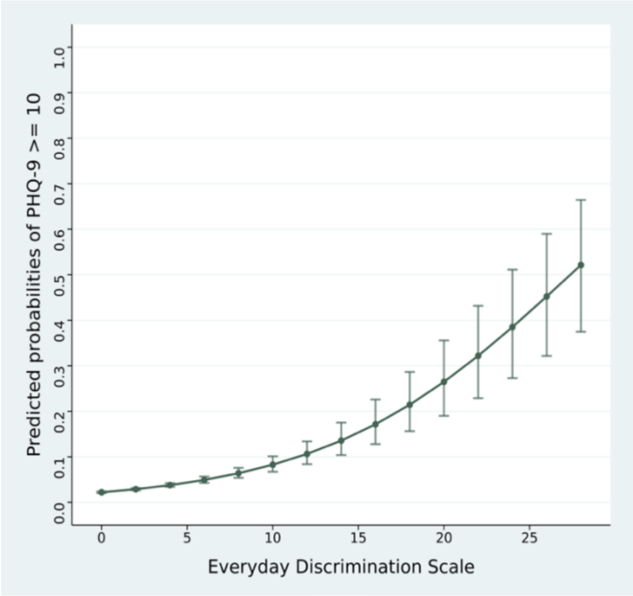
