## Supplemental Figure 4 for "Effect of everyday discrimination on depression and suicidal ideation during the COVID-19 pandemic: a large-scale, repeated-measures study in the *All of Us* Research Program"

**Supplemental Figure 4.** Variations in the effects of everyday discrimination on clinically elevated depression and suicidal ideation by pre-pandemic mood disorder diagnosis.

Outcome: Clinically elevated depression (PHQ-9 total score ≥ 10)

(a) Any reason(s)

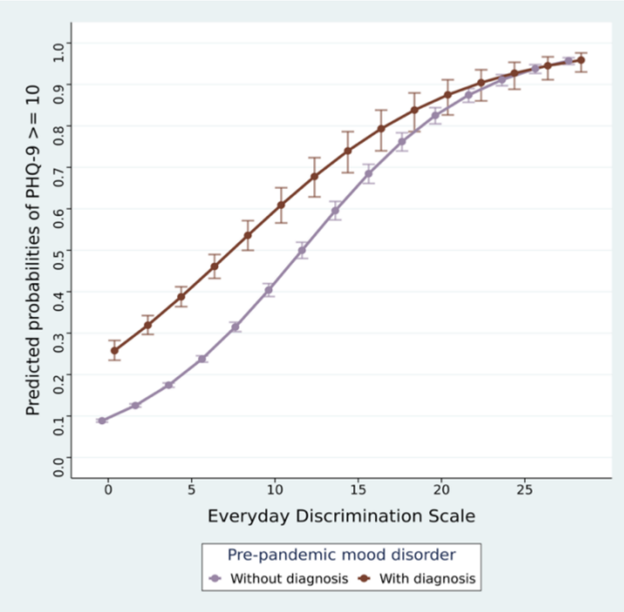


(b) Due to race or ancestry/national origin(s)

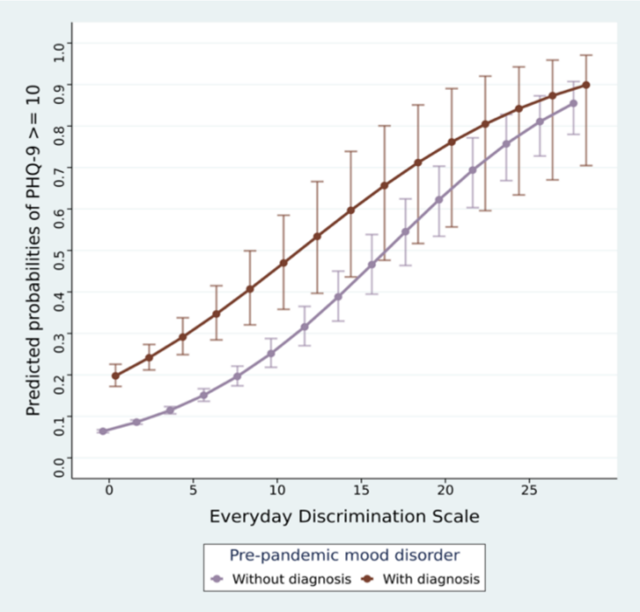


Outcome: Suicidal ideation (PHQ-9 item 9 > 0)

(c) Any reason(s)

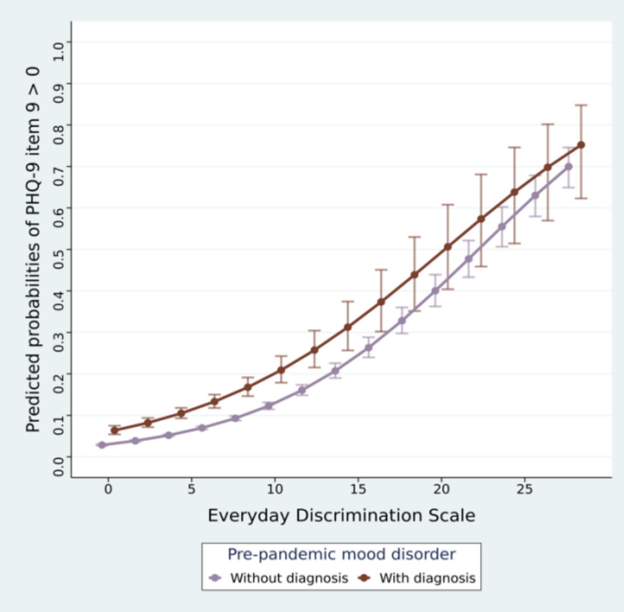


(d) Due to race or ancestry/national origin(s)

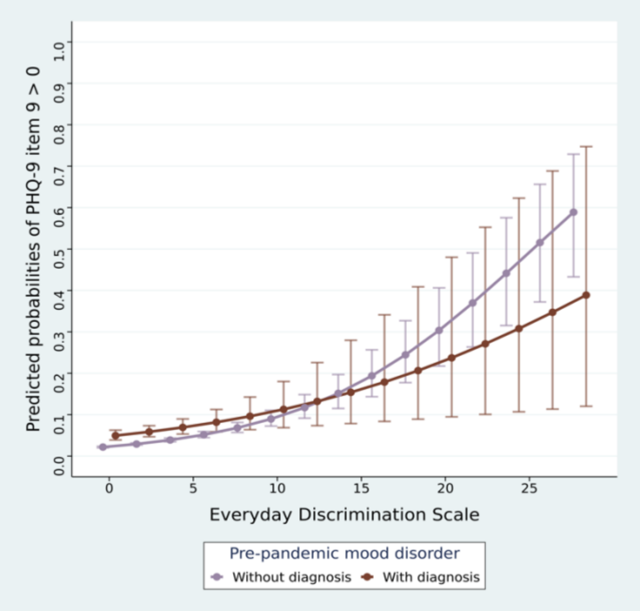
